# Occupational versus community risk of SARS-CoV-2 infection among employees of a long-term care facility: an observational study

**DOI:** 10.1101/2021.10.22.21265309

**Authors:** L. Lenggenhager, R. Martischang, J. Sauser, M. Perez, L. Vieux, C. Graf, S. Cordey, F. Laubscher, T. Robalo Nunes, Walter Zingg, A. Cori, S. Harbarth, M. Abbas

**Affiliations:** Infection Control Programme, Geneva University Hospitals, Geneva, Switzerland; Faculty of Medicine, University of Geneva, Geneva, Switzerland; Laboratory of Virology, Department of Diagnostics, Geneva University Hospitals, Geneva, Switzerland; MRC Centre for Global Infectious Disease Analysis, Imperial College London, London, United Kingdom; Occupational Health Service, Geneva University Hospitals, Geneva, Switzerland; Department of Rehabilitation and Geriatrics, Geneva University Hospitals, Geneva, Switzerland; Infectious Diseases Service of Hospital Garcia de Orta, EPE, Almada, Portugal

## Abstract

**Objectives:** We investigated the relative contribution of occupational (vs. community) exposure for severe acute respiratory syndrome coronavirus 2 (SARS-CoV-2) infection among employees of a university-affiliated long-term care facility (LTCF), during the 1^st^ pandemic wave in Switzerland (March to June 2020).

**Methods:** We performed a nested analysis of a seroprevalence study among all volunteering LTCF staff to determine community and nosocomial risk factors for SARS-CoV-2 seropositivity using modified Poison regression. We also combined epidemiological and genetic sequencing data from a coronavirus disease 2019 (COVID-19) outbreak investigation in a LTCF ward to infer transmission dynamics and acquisition routes of SARS-CoV-2, and evaluated strain relatedness using a maximum likelihood phylogenetic tree.

**Results:** Among 285 LTCF employees, 176 participated in the seroprevalence study, of whom 30 (17%) were seropositive for SARS-CoV-2. Most (141/176, 80%) were healthcare workers (HCWs). Risk factors for seropositivity included exposure to a COVID-19 inpatient (adjusted prevalence ratio [aPR] 2.6; 95%CI 0.9-8.1) and community contact with a COVID-19 case (aPR 1.7; 95%CI 0.8-3.5). Among 18 employees included in the outbreak investigation, the outbreak reconstruction suggests 4 likely importation events by HCWs with secondary transmissions to other HCWs and patients.

**Conclusions:** These two complementary epidemiologic and molecular approaches suggest a substantial contribution of both occupational and community exposures to COVID-19 risk among HCWs in LTCFs. These data may help to better assess the importance of occupational health hazards and related legal implications during the COVID-19 pandemic.

## Introduction

Long-term care facility (LTCF) healthcare workers (HCWs) are recognized vectors in the transmission chain between other HCWs and residents [1-3]. As most studies of transmission routes and risk factors for employee seroconversion were studied in acute-care settings [4, 5], evidence remains scarce for LTCFs, often weakened by flawed surveillance data [6].

In many countries, including Switzerland, COVID-19 among employees working in healthcare institutions caring for COVID-19 patients is automatically recognized as an occupational disease [7]. Nevertheless, the community contribution to SARS-CoV-2 acquisition among health sector employees and in nosocomial outbreaks has recently increasingly been pointed out [1, 4, 7-12]. To our knowledge, the exact contribution of community exposures versus occupational health hazards leading to SARS-CoV-2 infection among HCWs in LTCF or nursing home settings has not yet been determined.

Geneva University Hospitals (HUG) affiliated LTCFs are particularly befitted to estimate SARS-CoV-2 acquisition modes and infection rates among employees given the implementation of a robust surveillance system during the first pandemic wave. This includes (1) an institution-wide seroprevalence survey, (2) an outbreak investigation among LTCF patients and HCWs, (3) systematic, syndromic surveillance of employees, and (4) the systematic storage of viral isolates by the National Center of Emerging Viral Diseases hosted at HUG. Here we combine these epidemiologic, molecular, serological and genotypic data collected in the same LTCF to understand the relative contribution of occupational (versus community) exposure for COVID-19 infection among employees of a university-affiliated LTCF in Switzerland.

## Methods

### Study design and population

In this cohort study conducted from March 1 to June 30, 2020, in a HUG-affiliated LTCF, we combined data from 1) a previously published institution-wide prospective seroprevalence study in employees [13], 2) an outbreak investigation among HCWs and patients, and 3) all SARS-CoV-2 RT-PCR results performed in the context of syndromic surveillance among HUG employees.

We included a subgroup of LTCF employees among the 3’241 volunteering participants of the seroprevalence study [13]. The outbreak in one ward of the LTCF spanned from March 15 to April 8 (based on swab dates) and involved 12 patients (all nosocomial cases) and 23 HCWs, with 10 and 18 specimens analysed by whole genome sequencing (WGS), respectively. HCWs working in the LTCF with a positive SARS-CoV-2 RT-PCR and patients with nosocomial SARS-CoV-2 acquisition were included in the outbreak analysis. Finally, all LTCF employees who underwent SARS-CoV-2 RT-PCR testing at HUG were also included (see Supplements eAppendix 1 for details).

The seroprevalence survey and the outbreak investigation were approved by the SNF. Each study participant of the seroprevalence survey provided written informed consent.

### Study setting

HUG is the largest tertiary-care centre in Switzerland with >2000 beds and roughly 13’600 employees. It includes 8 campuses, 4 of which are part of the Department of Rehabilitation and Geriatrics and includes several LTCFs. Our study focuses on a LTCF which includes 8 wards, 4 dedicated to rehabilitation (104 beds) and 4 dedicated to patients awaiting nursing home placement (112 beds), with 285 employees.

### Outcomes and definitions

The primary outcome was SARS-CoV-2 seropositivity among employees, with community vs occupational risk factors as primary exposures of interest. Secondary outcomes included 1) transmission pathways (reconstructing the outbreak and determining who infected whom) and 2) the cumulative number of positive SARS-CoV-2 RT-PCRs among LTCF employees.

Occupational risk factors were classified as any exposure that may result from the performance of an employee’s duties (see Supplements eAppendix 1 and Table 1). Regarding the outbreak reconstruction, patients with nosocomial Covid-19 were included if they had a positive SARS-CoV-2 RT-PCR and onset of symptoms ≥ 5 days after admission in the LTCF, in accordance with Swissnoso guidelines [14]. This study was performed in accordance with the STROBE statement for cohort studies [15] and the ORION guidelines [16].

**Table 1.**
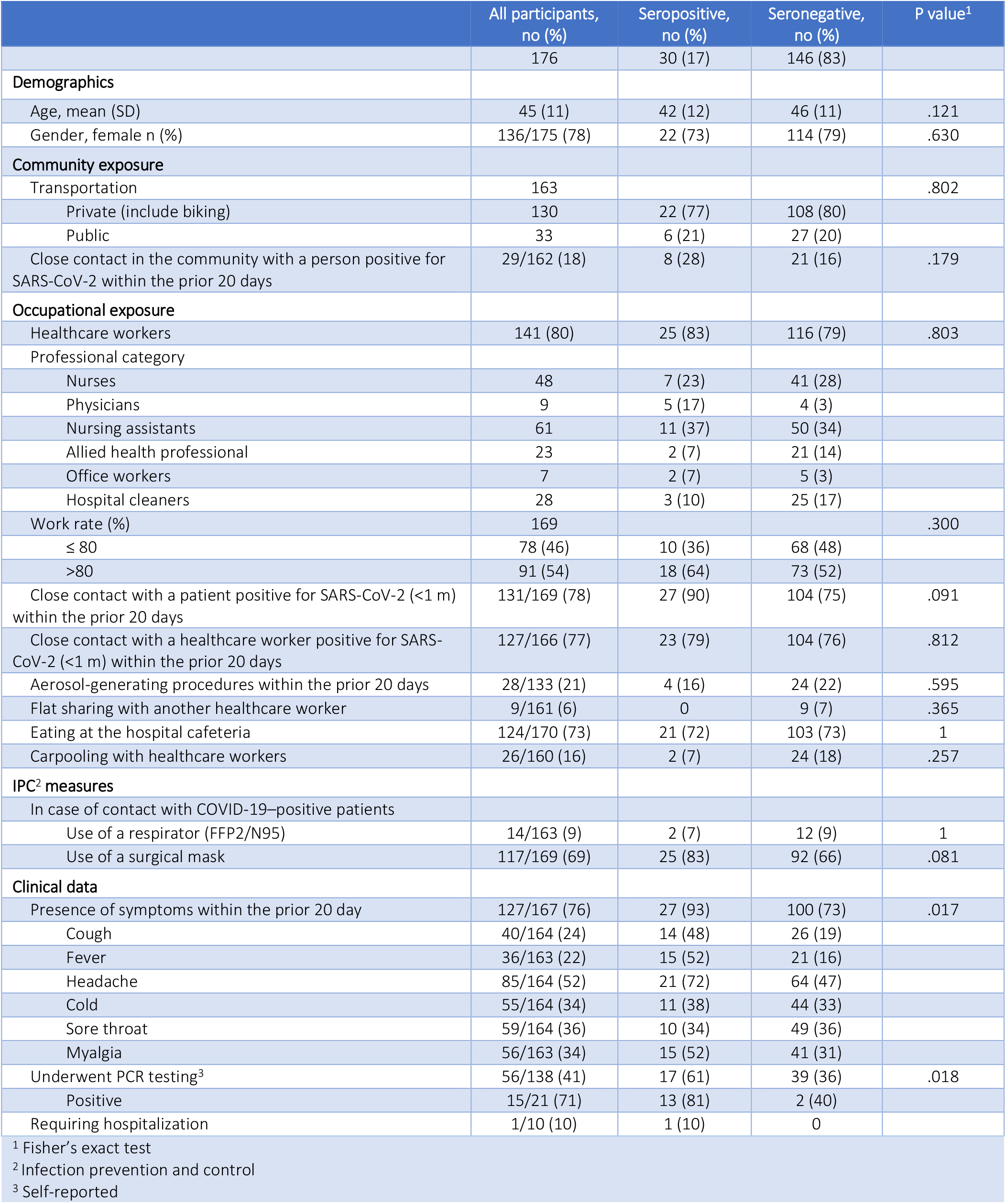
Clinical and demographic characteristics of LTCF employees participating in the seroprevalence study.

### Data sources

We retrieved LTCF employee data from the institution-wide prospective seroprevalence study [13]. We retrieved data from patients and HCWs included in the outbreak (dates of symptoms onset, date of positive SARS-CoV-2 RT-PCR) from a prospective national surveillance of all COVID-19 patients mandated by the Swiss Federal Office of Public Heath (FOPH) [17] and from the Department of Occupational Health, respectively. Data on SARS-CoV-2 RT-PCR results performed at HUG among LTCF employees from March 1 to June 30, 2020 were anonymously retrieved from the Department of Occupational Health surveillance database.

### Microbiological methods

As previously described, participants’ samples collected for the seroprevalence survey were analysed with a 2-tiered diagnostic strategy [13]. All COVID-19 cases included in the outbreak investigation were confirmed by RT-PCR on naso-pharyngeal swab and followed by WGS using an unbiased high-throughput sequencing method (see Supplements eAppendix 2 for details).

### Statistical analysis

We performed descriptive analyses with means (± standard deviations (SD)) or medians (interquartile range (IQR)) and proportions, as appropriate. For comparisons between groups we used Student’s *t*-test and Pearson’s chi-square test or Fisher’s exact test, for continuous and categorical variables respectively.

For the seroprevalence analysis, modified Poisson regression with robust variance was performed to determine and estimate risk factors for seropositivity. Variable selection using best subset regression with clinical consideration retained close contact (<1 m) with a COVID-19 inpatient within the previous 20 days and close contact with a laboratory-proven COVID-19 case in the community within the previous 20 days as independent variables (Supplements eAppendix 3). This model was compared to the null model using a likelihood ratio test and retained if it showed statistical significance.

As previously described [18], the outbreak reconstruction was performed combining epidemiological (date of symptom onset and trajectories) and genetic sequencing data. It includes 1) construction of an epidemic curve and estimation of the time-varying reproduction number (*Rt*) using the EpiEstim package [19], 2) an ancestry reconstruction (who infected whom) using the outbreaker2 package [20, 21], 3) a maximum posterior tree, 4) a maximum likelihood phylogenetic tree.

Analyses were performed with Stata v.15 (StataCorp, College Station, Texas, USA) and R version 4.0.4 (2021-02-15) (R Foundation for Statistical Computing, Vienna, Austria). Detailed methods are described in the Supplements eAppendix 3.

## Results

### Seroprevalence analysis

Among 285 employees working in the LTCF from March 1 to June 30, 2020, 199 (70%) participated in the seroprevalence survey and 176 (62%) were included in our analysis (see Supplements eAppendix 1 and Supplementary Figure 3). Thirty (17%) of the 176 participants were seropositive for SARS-CoV-2. The majority were female (136/175, 78%), HCWs with patient contact (141/176, 80%) and presented ≥1 symptom compatible with COVID-19 (127/167, 76%). Data regarding symptoms and gender were missing for 9 and 1 employees, respectively. Seropositive participants were more likely to be exposed to a COVID-19 case in the community (8/30, 28%) and to a COVID-19 patient at work (27/30, 90%) than seronegative participants, with 21/146 (16%) and 104/146 (75%) seronegative participants reporting a contact with a COVID-19 case in the community and with a COVID-19 patient at work, respectively. Clinical and demographic characteristics of employees are described in Table 1.

Multivariable analysis revealed that both occupational exposure (adjusted prevalence ratio aPR 2.6; 95%CI 0.9-8.1) and community exposure (aPR 1.7; 95%CI 0.8-3.5) were associated with higher risk of seropositivity, although these associations were not statistically significant.

When restricting the analysis to participants at baseline only (between April 8 and 16, 2020), including 20 (10%) seropositive employees and 179 (90%) seronegative employees, multivariable analysis revealed statistically significant associations between seropositivity and both occupational exposure (aPR 3.6; 95%CI 1.1-11.5, P=0.03) and community exposure (aPR 3.28; 95%CI 1.4-7.8, P=0.007).

### Outbreak investigation

The outbreak spanned from March 15 to April 8, 2020 based on swab dates, with 35 RT-PCR proven cases (12 patients and 23 HCWs). The attack rate was 50% and 23.5% for HCWs and patients, respectively. Figure 1A shows the epidemic curve based on dates of symptom onset. There was considerable uncertainty in the time-varying reproduction number (*Rt*); the first point estimate was 1.38 (95% credibility interval 0.82-2.10) on March 16, 2020, and decreased below 1 (0.89, 95%CrI 0.48-1.41) on March 20, 2020. (Figure 1B).

**Figure 1.**
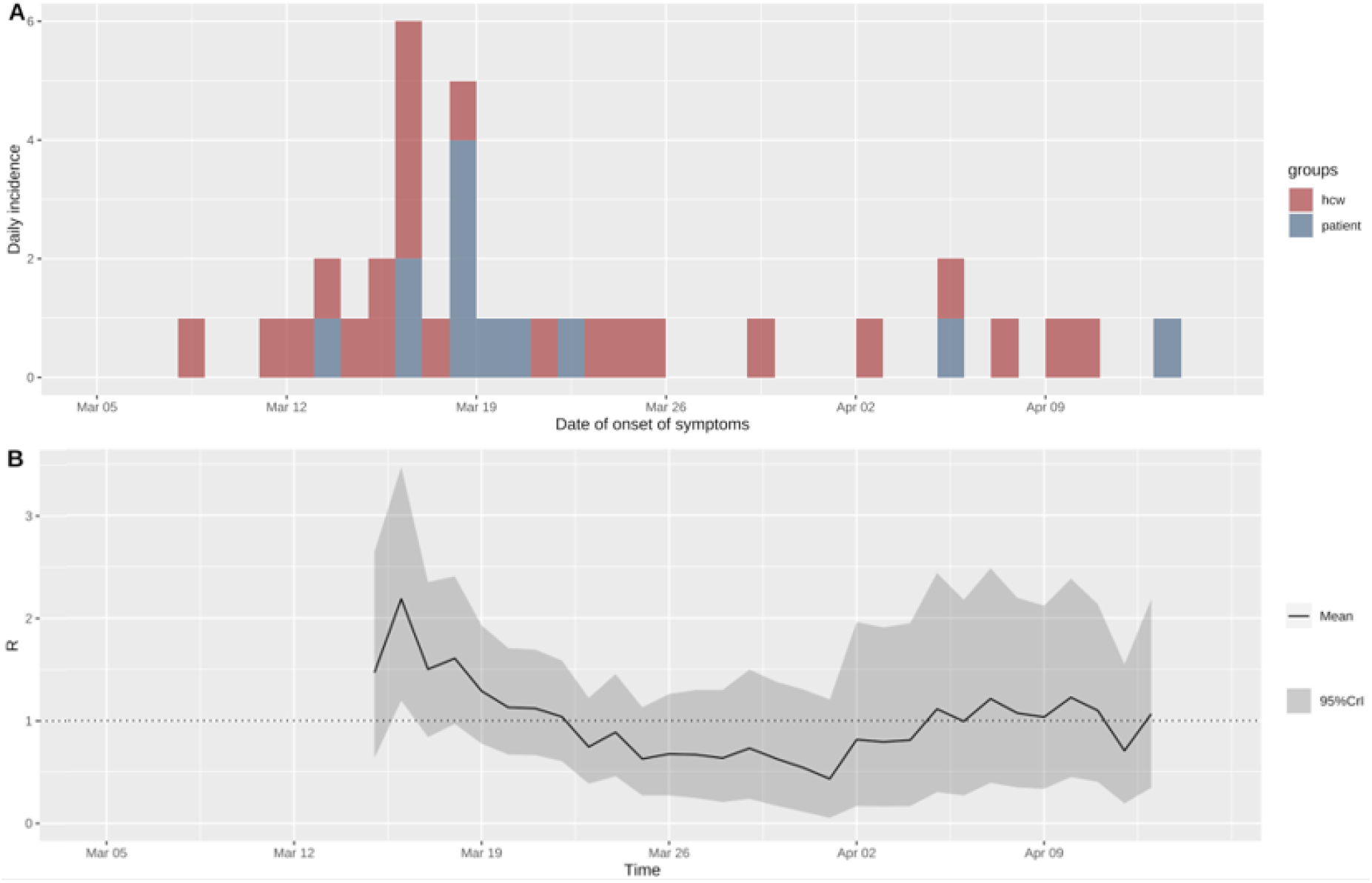
A. Epidemic curve of the nosocomial COVID-19 outbreak involving LTCF HCWs (red) and patients (blue). B. Estimated time-varying reproduction number (*Rt*) across the duration of the outbreak. EpiEstim provides *Rt* estimates on sliding weekly windows, i.e. the estimate of one day corresponds to the previous 7 days.

Genetic sequences were obtained and a phylogenetic tree was constructed for 18 HCWs (red) and 10 patients (blue) involved in the outbreak (Figure 2). We observed one large cluster of 10 HCWs and 10 patients (branch names G4504T, C10156T, and C14220T on Figure 2) with a sub-cluster including 3 HCWs and 2 patients (branch name G29703A on Figure 2), highly suggestive of nosocomial transmission. Given the sequences and their mutation similarities, it is reasonable to suggest that these 10 HCWs (56%), including one imported case, may have acquired SARS-CoV-2 in the course of their professional activities (clinical and non-clinical duties, commuting together, etc.). The 8 (44%) remaining strains retrieved from HCWs showed a significant amount of genetic diversity, and tend to cluster with community sequences.

**Figure 2.**
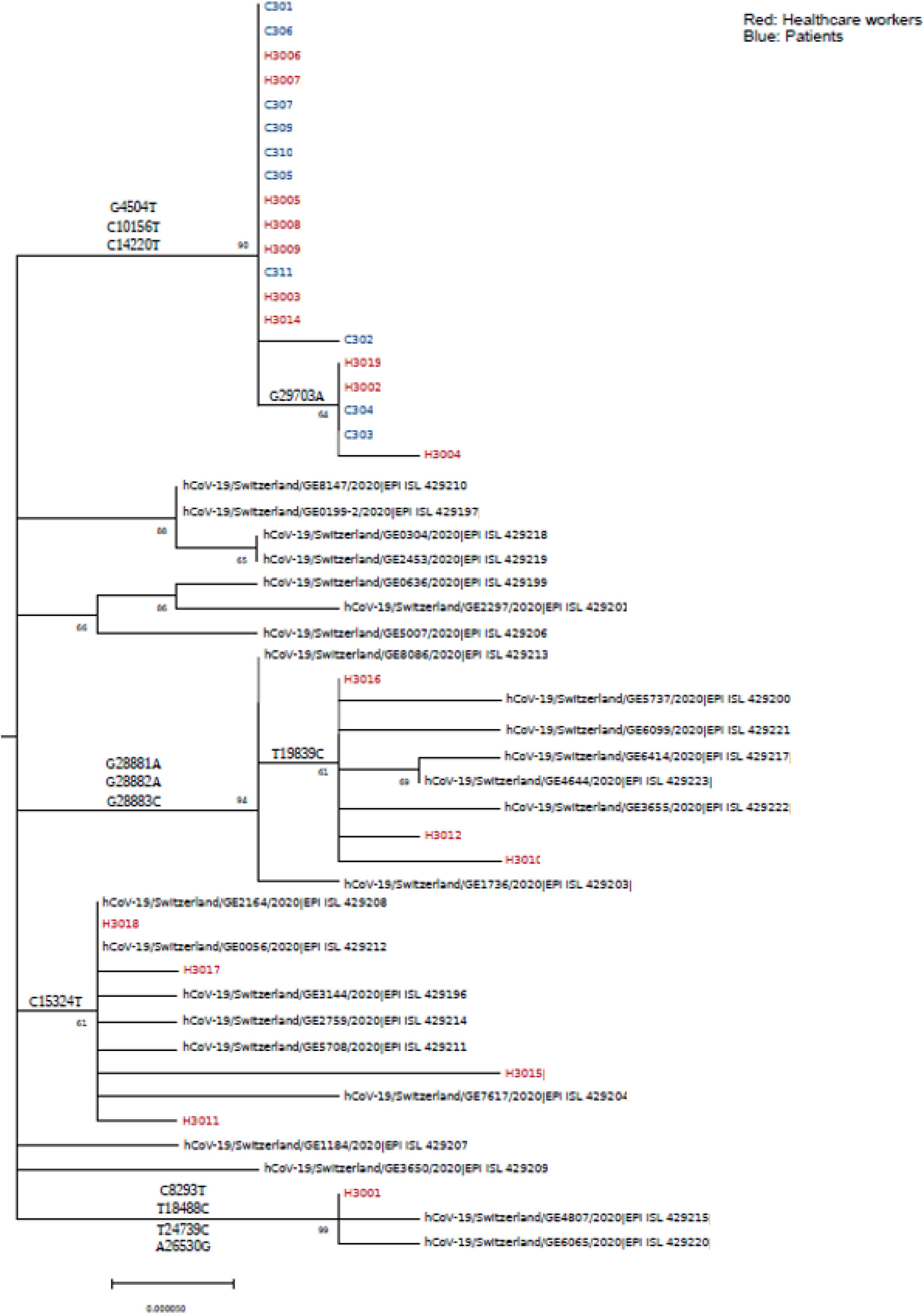
Maximum likelihood phylogenetic tree of SARS-CoV-2 genomes from 10 patients (blue) and 18 HCWs (red) sequenced during the outbreak investigation.

Of these 8 strains sharing similarity with community sequences, the ancestry reconstruction highlighted that 4 imported cases among HCWs (H3015, H3017, H3001 and H3016; Supplementary Figure 4) may have secondarily infected 3 other HCWs (H3010, H3012 and H3018), among which one (H3018) secondarily infected another HCWs (H3011), with an estimated posterior support of 100% (H3010, H3012 and H3011) and 58% (H3018) for these transmission links (Supplementary Figure 5). Thus, it seems that 4 of the 8 remaining strains from HCWs were infected by other HCWs carrying community sequences.

## Discussion

This comprehensive study of SARS-CoV-2 acquisition and transmission dynamics conducted in a Swiss LTCF during the 1^st^ pandemic wave shows that both community and occupational exposures play an important role leading to COVID-19 among employees. Overall, these complementary approaches challenge traditional assumptions that SARS-CoV-2 infection in hospital employees is always due to an occupational exposure. Moreover, our results show that the probable 4 imported cases in the initial outbreak were all HCWs harbouring a community sequence (H3015, H3017, H3001, H3016 on Figure 2, Supplementary Figure 4 and 5), who then secondarily infected their colleagues. Even if the outbreak analysis is unable to confirm that these transmissions between HCWs occurred at work or outside work, our study highlights the important contribution of HCWs and community importation in the transmission chain of this LTCF outbreak of COVID-19.

Our results underline the major role of community-acquired infection in LTCF HCWs, who may then introduce the virus into the facility, and are in line with most studies investigating the occupation health hazard of COVID-19 among hospital employees, even though the latter mostly focused on acute care settings [8, 10, 11, 13, 22-26].

Regarding occupational exposure, HCWs, and particularly LTCF HCWs, are known to be at higher risk for SARS-CoV-2 infection than the general population [27-31]. Although this consideration has led to huge efforts to protect HCWs and patients from nosocomial infection [7, 29, 32], this may have led to an overestimation of the importance of patient-to-HCW transmission compared to HCW-to-HCW or community-to-HCW transmission [2, 33].

Our results strongly emphasize the importance of HCW-to-HCWs SARS-CoV-2 transmission, as previously described [18, 34] and thus non only challenge the misperception that patients are the dominant reservoirs and vectors, but also support the importance of non-pharmaceutical interventions and repeated testing of employees to limit transmission from community to healthcare settings, as well as the importance of aggressive vaccination campaigns among LTCF employees [35-37]. Therefore, occupational exposure is not always related to direct patient care.

Disentangling the real occupational vs community health hazard of COVID-19 infection among LTCF employees is crucial given the proportion of long COVID syndrome reported in recent studies and the related legal and economic implications. Indeed, in the Swiss legal landscape, the sick leaves due to COVID-19 among employees in contact with patients would be covered by the Federal Law on Accident Insurance and Disability Insurance depending on the duration of the leave, in contrast to Health Insurance when the leave is due to non-occupational disease. Up to two-thirds of infected middle-aged individuals (40-60 years old) may indeed suffer from disabling symptoms potentially lasting for months and leading to long periods of sick leave for 11% of them [38-40].

The main strength of our study is that, to our knowledge, it is the first quantitative analysis of occupational vs community risk of COVID-19 among LTCF employees combining epidemiological, serological, and molecular data. Thus, these original results add precious information regarding SARS-CoV-2 spread in LTCFs, which share many similarities with nursing homes. This study also presents some limitations, including 1) potential selection bias regarding the fact that not all LTCF employees participated in the seroprevalence survey; 2) information and potential recall bias given the retrospective data collection; 3) the fact that contact with an asymptomatic COVID-19 case (community or hospital) may have not been recorded; 4) different populations in the two sets of analysis; 5) lack of power given the small sample size and number of events observed. Finally, despite a non-statistically significant result, and the possibility of residual confounding, the 95%CI of the influence of community on LTCF employees seropositivity clearly indicates a probable exposure effect, which is supported by seroprevalence survey at baseline and detailed genomic analysis.

In conclusion, these two complementary approaches demonstrate a substantial contribution of both occupational and community exposures to seropositivity and infection risk. The role of HCWs in preventing importation of SARS-CoV-2 to LTCFs from the community is crucial. These data may not only allow to better assess occupational health hazards and related legal implications during and after the COVID-19 pandemic, but also emphasize the urgent need to maximise vaccine uptake in LTCF HCWs in order to limit HCW-to-HCW and HCW-to-patient transmission.

## Data Availability

For confidentiality reasons, data are not available.

## Acknowledgements

The authors would like to thank Dr. Catherine Chenaud (Occupational Health Department of the Geneva University Hospital) and Mr. Jean-Claude Suard (Occupational Health Department of the Geneva University Hospital) for providing us with employees RT-PCR data and all employees from the virological laboratory of the Geneva University Hospital for their work.

Anne Cori is supported by the National Institute for Health Research (NIHR) Health Protection Research Unit in Modelling and Health Economics, a partnership between Public Health England, Imperial College London and LSHTM (grant code NIHR200908); and acknowledges funding from the MRC Centre for Global Infectious Disease Analysis (reference MR/R015600/1), jointly funded by the UK Medical Research Council (MRC) and the UK Foreign, Commonwealth & Development Office (FCDO), under the MRC/FCDO Concordat agreement and is also part of the EDCTP2 programme supported by the European Union.

## Disclaimer

“The views expressed are those of the author(s) and not necessarily those of the NIHR, Public Health England or the Department of Health and Social Care.”

## References

1. Sikkema RS PS, Nieuwenhuijse DF, O’Toole Á, Verweij J, van der Linden A, et al. COVID-19 in health-care workers in three hospitals in the south of the Netherlands: a cross-sectional study. Lancet Infect Dis. 2020 Nov;(11):1273–80.

2. Abbas M, Robalo Nunes T, Martischang R, Zingg W, Iten A, Pittet D, et al. Nosocomial transmission and outbreaks of coronavirus disease 2019: the need to protect both patients and healthcare workers. Antimicrob Resist Infect Control. 2021;10(1):7.

3. MacCannell T, Batson J, Bonin B, Kc A, Quenelle R, Strong B, et al. Genomic epidemiology and transmission dynamics of SARS-CoV-2 in congregate healthcare facilities in Santa Clara County, California. Clin Infect Dis. 2021;Online ahead of print.

4. Braun KM, Moreno GK, Buys A, Somsen ED, Bobholz M, Accola MA, et al. Viral sequencing reveals US healthcare personnel rarely become infected with SARS-CoV-2 through patient contact. Clin Infect Dis. 2021;73(6):e1329–e36.

5. Nguyen LH, Drew DA, Joshi AD, Guo CG, Ma W, Mehta RS, et al. Risk of COVID-19 among frontline healthcare workers and the general community: a prospective cohort study. medRxiv. 2020;Preprint. 2020 May 25.

6. Arons MM, Hatfield KM, Reddy SC, Kimball A, James A, Jacobs JR, et al. Presymptomatic SARS-CoV-2 Infections and Transmission in a Skilled Nursing Facility. N Engl J Med. 2020;382(22):2081–90.

7. Carlsten C, Gulati M, Hines S, Rose C, Scott K, Tarlo SM, et al. COVID-19 as an occupational disease. Am J Ind Med. 2021;64(4):227–37.

8. Kahlert CR, Persi R, Gusewell S, Egger T, Leal-Neto OB, Sumer J, et al. Non-occupational and occupational factors associated with specific SARS-CoV-2 antibodies among hospital workers - A multicentre cross-sectional study. Clin Microbiol Infect. 2021;27(9):1336–44.

9. Schneider S, Piening B, Nouri-Pasovsky PA, Kruger AC, Gastmeier P, Aghdassi SJS. SARS-Coronavirus-2 cases in healthcare workers may not regularly originate from patient care: lessons from a university hospital on the underestimated risk of healthcare worker to healthcare worker transmission. Antimicrob Resist Infect Control. 2020;9(1):192.

10. Baker JM, Nelson KN, Overton E, Lopman BA, Lash TL, Photakis M, et al. Quantification of Occupational and Community Risk Factors for SARS-CoV-2 Seropositivity Among Health Care Workers in a Large U.S. Health Care System. Ann Intern Med. 2021;174(5):649–54.

11. Jacob JT, Baker JM, Fridkin SK, Lopman BA, Steinberg JP, Christenson RH, et al. Risk Factors Associated With SARS-CoV-2 Seropositivity Among US Health Care Personnel. JAMA Netw Open. 2021;4(3):e211283.

12. Chun-Wai Wong R, Lee MK, Kit-Hang Siu G, Lee LK, Siu-Lun Leung J, Chi-Man Leung E, et al. Healthcare workers acquired COVID-19 disease from patients? An investigation by phylogenomics. J Hosp Infect. 2021;115:59–63.

13. Martischang R, Iten A, Arm I, Abbas M, Meyer B, Yerly S, et al. Severe acute respiratory coronavirus virus 2 (SARS-CoV-2) seroconversion and occupational exposure of employees at a Swiss university hospital: A large longitudinal cohort study. Infect Control Hosp Epidemiol. 2021:1–8.

14. Swissnoso. Prevention & control of healthcare-associated COVID-19 outbreaks https://www.swissnoso.ch/fileadmin/swissnoso/Dokumente/5_Forschung_und_Entwicklung/6_Aktuelle_Erreignisse/200515_Prevention_and_control_of_healthcare-associated_COVID-19_outbreaks_V1.0_ENG.pdf 2020 [

15. STROBE Checklist for cohort studies https://www.strobe-statement.org/fileadmin/Strobe/uploads/checklists/STROBE_checklist_v4_cohort.pdf [

16. ORION Checklist of items to include when reporting an outbreak or intervention study of a nosocomial organism https://www.ucl.ac.uk/drupal/site_antimicrobial-resistance/sites/antimicrobial-resistance/files/checklist_authors.pdf.

17. Thiabaud A, Iten A, Balmelli C, Senn L, Troillet N, Widmer A, et al. Cohort profile: SARS-CoV-2/COVID-19 hospitalised patients in Switzerland. Swiss Med Wkly. 2021;151:w20475.

18. Abbas M, Nunes TR, Cori A, Cordey S, Laubscher F, Baggio S, et al. Explosive nosocomial outbreak of SARS-CoV-2 in a rehabilitation clinic: the limits of genomics for outbreak reconstruction. J Hosp Infect. 2021;Article in press.

19. Cori A, Ferguson NM, Fraser C, Cauchemez S. A new framework and software to estimate time-varying reproduction numbers during epidemics. Am J Epidemiol. 2013;178(9):1505–12.

20. Jombart T, Cori A, Didelot X, Cauchemez S, Fraser C, Ferguson N. Bayesian reconstruction of disease outbreaks by combining epidemiologic and genomic data. PLoS Comput Biol. 2014;10(1):e1003457.

21. Campbell F, Strang C, Ferguson N, Cori A, Jombart T. When are pathogen genome sequences informative of transmission events? PLoS Pathog. 2018;14(2):e1006885.

22. Eyre DW, Lumley SF, O’Donnell D, Campbell M, Sims E, Lawson E, et al. Differential occupational risks to healthcare workers from SARS-CoV-2 observed during a prospective observational study. Elife. 2020;9.

23. Nygren D, Noren J, De Marinis Y, Holmberg A, Fraenkel CJ, Rasmussen M. Association between SARS-CoV-2 and exposure risks in health care workers and university employees - a cross-sectional study. Infect Dis (Lond). 2021;53(6):460–8.

24. Steensels D, Oris E, Coninx L, Nuyens D, Delforge ML, Vermeersch P, et al. Hospital-Wide SARS-CoV-2 Antibody Screening in 3056 Staff in a Tertiary Center in Belgium. JAMA. 2020;324(2):195–7.

25. Galanis P, Vraka I, Fragkou D, Bilali A, Kaitelidou D. Seroprevalence of SARS-CoV-2 antibodies and associated factors in healthcare workers: a systematic review and meta-analysis. J Hosp Infect. 2021;108:120–34.

26. Candel FJ, Barreiro P, San Roman J, Del Mar Carretero M, Sanz JC, Perez-Abeledo M, et al. The demography and characteristics of SARS-CoV-2 seropositive residents and staff of nursing homes for older adults in the Community of Madrid: the SeroSOS study. Age Ageing. 2021;50(4):1038–47.

27. Iversen K, Bundgaard H, Hasselbalch RB, Kristensen JH, Nielsen PB, Pries-Heje M, et al. Risk of COVID-19 in health-care workers in Denmark: an observational cohort study. The Lancet Infectious Diseases. 2020;20(12):1401–8.

28. Gomez-Ochoa SA, Franco OH, Rojas LZ, Raguindin PF, Roa-Diaz ZM, Wyssmann BM, et al. COVID-19 in Health-Care Workers: A Living Systematic Review and Meta-Analysis of Prevalence, Risk Factors, Clinical Characteristics, and Outcomes. Am J Epidemiol. 2021;190(1):161–75.

29. Piccoli L, Ferrari P, Piumatti G, Jovic S, Rodriguez BF, Mele F, et al. Risk assessment and seroprevalence of SARS-CoV-2 infection in healthcare workers of COVID-19 and non-COVID-19 hospitals in Southern Switzerland. Lancet Reg Health Eur. 2021;1:100013.

30. Akinbami LJ, Chan PA, Vuong N, Sami S, Lewis D, Sheridan PE, et al. Severe Acute Respiratory Syndrome Coronavirus 2 Seropositivity among Healthcare Personnel in Hospitals and Nursing Homes, Rhode Island, USA, July-August 2020. Emerg Infect Dis. 2021;27(3):823–34.

31. Stringhini Sea. Seroprevalence of anti-SARS-CoV-2 IgG antibodies in Geneva, Switzerland (SEROCoV-POP): a population-based study. Lancet. 2020;396(10247):313–9.

32. Verbeek JH, Rajamaki B, Ijaz S, Sauni R, Toomey E, Blackwood B, et al. Personal protective equipment for preventing highly infectious diseases due to exposure to contaminated body fluids in healthcare staff. Cochrane Database Syst Rev. 2020;4:CD011621.

33. Mylle G, Vanacker H, Verbeek C, Bulterys S, Godderis L, De Schryver A. Prevalence of SARS-CoV-2 among Belgian workers in long-term care facilities. Occup Med (Lond). 2021; Online ahead of print.

34. Robalo Nunes T, Lebowitz D, Fraccaro M, Perez M, Vieux L, Abbas M, et al. Can long-term care facilities remain a coronavirus disease 2019 (COVID-19)-free bubble? An outbreak report. Infect Control Hosp Epidemiol. 2021:1–2.

35. Moghadas SM, Vilches TN, Zhang K, Wells CR, Shoukat A, Singer BH, et al. The impact of vaccination on COVID-19 outbreaks in the United States. Clin Infect Dis. 2021; Online ahead of print.

36. Holmdahl I, Kahn R, Hay JA, Buckee CO, Mina MJ. Estimation of Transmission of COVID-19 in Simulated Nursing Homes With Frequent Testing and Immunity-Based Staffing. JAMA Netw Open. 2021;4(5):e2110071.

37. Evans S, Agnew E, Vynnycky E, Stimson J, Bhattacharya A, Rooney C, et al. The impact of testing and infection prevention and control strategies on within-hospital transmission dynamics of COVID-19 in English hospitals. Philos Trans R Soc Lond B Biol Sci. 2021;376(1829):20200268.

38. Yelin D, Margalit I, Yahav D, Runold M, Bruchfeld J. Long COVID-19-it’s not over until? Clin Microbiol Infect. 2021;27(4):506–8.

39. Carvalho-Schneider C, Laurent E, Lemaignen A, Beaufils E, Bourbao-Tournois C, Laribi S, et al. Follow-up of adults with noncritical COVID-19 two months after symptom onset. Clin Microbiol Infect. 2021;27(2):258–63.

40. Xiong Q, Xu M, Li J, Liu Y, Zhang J, Xu Y, et al. Clinical sequelae of COVID-19 survivors in Wuhan, China: a single-centre longitudinal study. Clin Microbiol Infect. 2021;27(1):89–95.

