## Supplementary Appendix for "Occupational versus community risk of SARS-CoV-2 infection among employees of a long-term care facility: an observational study": Supplementary_Appendix.docx

This appendix has been provided by the authors to give readers additional information about their work.

### **eAppendix 1**. Data source, participants and definitions

#### Seroprevalence survey

As previously described [1], All HUG volunteering employees (n= 3,421) were recruited by study nurses who visited wards and office areas. The recruitment period occurred from March 30, 2020, to April 17, 2020, with 2 follow-up periods from April 17 to May 11 and from May 11 to June 12. More precisely, the recruitment period, or visit 1 (V1), for employees working in the LTCF occurred at baseline from April 8 to 16, with 2 follow-up, or visit 2 (V2) and 3 (V3), period from April 29 and May 7 and from May 20 to 28. Each participant was asked to attend every 3 weeks. At each visit, a serum sample was drawn, and health-related data were collected using a self-completed questionnaire under the supervision of medical students. Employees not able to provide serum sample and/or questionnaire were excluded from the visit but could participate in the other visit.

Among all participants included in the seroprevalence survey, we selected 199 employees working in the LTCF. We performed descriptive analysis at baseline and at the last visit (V3). At baseline, 20/199 (10%) employees were seropositive instead of 30/176 (17%) when considered at V3. Thus, we decided to examine the status of seropositivity and exposures at V3 in order to increase the sample size of seropositive participants.

Among the 199 participants at baseline, 20 completed one visit, 21 completed two visits, and 158 completed have all three visits. Considering the participants with only one of the 3 visits completed, we included those who (1) are seropositive at the visit attended (3 participants) and (2) are seronegative at V3 (1 participants). The participants attending one visit and seronegative at the visit V1 or V2 were excluded because of an unknown serological status at V3 (16 participants excluded). Considering the participants with two visits completed we included those who (1) had one of the two visits attended at V3 (13 participants) and (2) attended V1 and V2 but were seropositive at one of the two visits (1 participant). Considering the employees participating to the three visits, they were all included (158 participants). Finally, 176 participants were included in our analysis (Supplementary Figure 3).

Risks factors for seroconversion were classified as community exposure (close contact with SARS-CoV-2–positive persons within or outside the household within the prior 20 days, number of household members, use of public versus private transportation) and nosocomial exposure (aerosol-generating procedures within the prior 20 days, close contact of <1 m with a COVID-19 patient within the prior 20 days, professional category, work rate, close contact of < 1 m with a SARS-CoV-2 positive healthcare worker within the prior 20 days, eating at the hospital cafeteria, carpooling with healthcare workers).

#### SARS-CoV-2 RT-PCR of employees

We anonymously collected all SARS-CoV-2 RT-PCR results of symptom based testing realized among symptomatic LTCF employees nasopharyngeal swab from March 1 2020 to June 30 2020 from the Department of Occupational Health. We did not perform regular screening of asymptomatic employees except in the context of outbreaks. Gender, age and profession were demographic data available in their database.

### **eAppendix 2**. Microbiological methods

All COVID-19 cases were confirmed by RT-PCR on naso-pharyngeal swab using the Cobas 6800 SARS CoV2 RT-PCR (Roche, Switzerland), the BD SARS-CoV2 reagent kit for BD Max system (Becton, Dickinson and Co, USA), and an in-house method based on the Charité assay [2].

#### Serological testing of anti-SARS-CoV-2 IgG antibodies

The two-step strategy comprises the use of a first S1 protein based IgG enzyme linked immunosorbent assay (ELISA; EUROIMMUN AG, Lübeck, Germany, no. EI 2606-9601 G) followed, in case of undetermined or positive results, by an ElectroChemiLuminescence ImmunoAssay (ECLIA) Elecsys Anti-SARS-CoV-2 Ig (Roche Diagnostics, Germany) according to the manufacturer’s instructions[1].

Unbiased high-throughput sequencing (HTS) analysis
As previously described [3], all nasopharyngeal swabs (NPS) with cycle threshold (Ct) values ≤ 30 or < 35 obtained for the E gene (Cobas 6800 SARS CoV2 RT-PCR and the Charite assays) and the S gene (BD SARS-CoV2 reagent kit for BD Max system), respectively, and for which sufficient volume remained, were selected for unbiased HTS analysis (n=35). HTS analysis was performed using the RNA protocol previously published [4]. Libraries were loaded on the HiSeq 4000 platform (Illumina) using the 2x100-bp protocol with dual indexing. Duplicate reads were removed using cd-hit (v4.6.8). Low-quality and adapter sequences were trimmed out using Trimmomatic (v0.33). Reads were then mapped against the reference sequence MN908947 using snap-aligner (v1.0beta.18). Consensus for sequences with at least 10-fold coverage were then generated using custom script. In parallel, all raw data were analyzed using a bioinformatic pipeline [5] that used virusscan 1.0 (https://github.com/sib-swiss/virusscan) to map reads against a respiratory viruses restricted database from Virosaurus [6] for the detection of co-infections. NPS with either Ct values > 30 or ≥ 35 obtained for the E gene (Cobas 6800 SARS CoV2 RT-PCR and the Charite assays) and the S gene (BD SARS-CoV2 reagent kit for BD Max system), respectively, or for which low SARS-CoV-2 genome coverage were obtained (i.e. < 80%) by the unbiased HTS method, were sequenced with an amplicon-based sequencing method if a sufficient volume remained. Thus, nucleic acids were extracted using the NucliSENS easyMAG (bioMérieux, Geneva, Switzerland) and them sequenced using an updated version of the nCoV-2019 sequencing protocol (https://www.protocols.io/view/ncov-2019-sequencing-protocol-bbmuik6w) (Microsynth, Balgach, Switzerland) on a MiSeq instrument (Illumina) with a 2x250-bp protocol.

#### Phylogenetic analysis

Sequence alignment was performed with MUSCLE (v3.8.31). The Evolutionary analyses were conducted in MEGA X [7] using the Maximum Likelihood method and Tamura 3-parameter model [8]. The phylogenetic tree includes also all SARS-CoV-2 complete genomes sequenced by our laboratory and submitted to GISAID from respiratory samples from COVID-19 positive patients presenting to our institute or others medical center in Geneva, Switzerland, during the same period [3].

### **eAppendix 3**. Statistical analysis

#### Seroprevalence survey: multivariable analysis

##### Variable selection

The variables selected for adjustment in multiple regression was based on clinical relevance; it included age, gender, professional category, close contact of <1 m with a COVID-19 patient within the prior 20 days, close contact of < 1 m with a SARS-CoV-2 positive healthcare worker within the prior 20 days, work rate (<=80%, >80%), use of surgical masks or use of respirators (FFP2/N95) In case of contact with COVID-19–positive patients, eating at the hospital cafeteria, close contact with SARS-CoV-2–positive persons within or outside the household within the prior 20 days, use of public versus private transportation, number of household members.

We considered best subset selection using these variables. The 9 best models according to AIC included either “close contact with SARS-CoV-2–positive persons within or outside the household within the prior 20 days”, “Use of surgical masks”, “work rate” or “close contact of <1 m with a COVID-19 patient within the prior 20 days”. Given the limited number of events (30 seropositive cases), using the 1:10 rule of thumb, we considered models using at most 3 variables. For information, only one model, the 7th best, included the 4 variables.

“Close contact with SARS-CoV-2–positive persons within or outside the household within the prior 20 days” and “close contact of <1 m with a COVID-19 patient within the prior 20 days” were of most interest. Likelihood ratio test comparing the model including these two variables to the null model showed statistical significance. In a second step, the same tests were applied to test the addition of a third variable, either “use of surgical masks” or “work rate”. None of these tests showed statistical significance. Hence, the model including “Close contact with SARS-CoV-2–positive persons within or outside the household within the prior 20 days” and “close contact of <1 m with a COVID-19 patient within the prior 20 days” was retained as the final model.

“Surgical masks” appeared in 8 of the 9 best models. The question asked was: “Did you use surgical masks in the past 20 days while in contact with patients?” However, subjects without contact with patients also replied to this question. We were thus unsure of the “quality” of the answers and so did not include this variable as a primary risk factor.

##### Modified Poisson regression

We uses the so-called “modified Poisson regression” which when applied to binary data provides estimates of prevalence ratio which are easier to understand and communicate [9]. The “modified” stand for the use of a robust variance, because when applying Poisson regression to binary data, the estimated variance is overestimated.

#### Outbreak reconstruction

We used the dates of symptoms onset to produce an epidemic curve; we inferred the date of onset using the median observed delay between symptoms onset and nasopharyngeal swab when the onset of symptoms was unknown.

We used the outbreaker2 R package [10-12] to reconstruct the outbreak by combining epidemiological and genetic data, as previously performed for SARS-CoV-2 [3]. Briefly, the model uses a Bayesian framework, which combines information on the generation time (time between infections in an infector/infected pair), contact patterns, with a model of sequence evolution to probabilistically reconstruct the transmission tree. Outbreaker2 runs a pre-specified number Markov-chain Monte Carlo iterations, from which 1000 are retained; each of these iterations represents a posterior transmission tree.

We ran a model using the following parameters:

- Mean serial interval of 3.0 days (standard deviation 4.1 days) [13]
- Mean incubation period of 5.95 days, (standard deviation 4.31 days) [14]
- 500’000 Markov-chain Monte Carlo iterations, with a thinning of 1 in 500, a burn-in of 500
- Uniform prior between 0.55 and 1 for the probability of a case being reported ("pi")
- Maximum "kappa" of 3 (i.e. we allowed for up to 2 unobserved cases on a transmission chain between any two observed cases)
- Default prior for the mutation rate ("mu") (uninformative exponential prior with mean 1)
- Where relevant, default prior for the non-infectious contact rate ("lambda") and contact reporting coverage ("eps"), i.e. uniform on [ 0, 1 ]
- Default likelihoods for all models, except for the model without contact data where this was disabled

We used the outbreaker2 package, which is designed to use contact tracing data to inform who infected whom. These data were unavailable for our outbreak, but we made a series of assumptions, as in a previous paper [3] to generate a matrix of possible contacts between cases. Patient administrative data were used to determine their presence in the ward. Because of potential errors in the shift data, we chose to make the conservative assumption that HCWs were present in the ward every day until the date of their first positive swab (included). Sensitivity analysis were performed without the contact data, which is the most conservative as the model makes no assumption about who may infect whom (i.e. all cases have an equal probability of infecting other cases).

In order to determine which cases are imported, in a first run of the model, outbreaker2 evaluates the global influence of each case based on their genetic sequences. By default, cases with a global influence 5 times higher than the average global influence are considered to be imported. Although the value of this threshold is set arbitrarily, it has excellent specificity.

Considering the large degree of homogeneity among the sequences used in this study, we lowered the default threshold set at 5 to a value of 3, in order to increase sensitivity of detection of imported cases. We performed sensitivity analyses for values of the threshold of 2, 4, and 5.

We assessed convergence of the model through visual inspection of the traces, and through the Gelman-Rubin convergence diagnostic (using the gelman.diag function in the R package coda v0.19-4) [15].

### **eAppendix 4**. Supplementary results

#### Syndromic RT-PCR testing among LTCF employees

Among the 285 LTCF employees, 106 (37%) had 125 RT-PCR tests on nasopharyngeal swabs and 18 (6%) were involved in the outbreak. Of all employees with screening based on syndromic surveillance, 36 (34%) presented at least one positive SARS-CoV-2 RT-PCR (Supplementary Table 1, Supplementary Figures 1 and 2). Forty-one (23%) of the 176 participants underwent SARS-CoV-2 RT-PCR testing at HUG, including 13 positive (32%) and 28 negative (68%) results (Supplementary Figure 1). Thirteen (43%) of the seroconverted participants had a positive SARS-CoV-2 RT-PCR compared to 26 (18%) in the seronegative group (n=146).

#### Outbreak reconstruction

The model converged well according to visual inspection, and with upper limits of the 95% confidence interval of the Gelman-Rubin diagnostic test <1.1. Due to the dense nature of the outbreak, and because it occurred in a single ward with a high degree of mixing, we did not observe significant differences between the model with contact data and the model without contact data (not shown).

##### Number of secondary infection per type of case

We calculated the number of secondary (i.e. onwards) infections for each case as summed over all posterior trees (Supplementary Figure 6). This represents the individual reproductive number (R), which we stratified by type of case (i.e. patient vs HCW). We assessed whether HCWs and patients had similar proportions of at least 1 onward case using the chi-squared test, and whether the cumulative distribution of secondary cases differed between type of case using the Kolmogorov-Smirnov test.

HCWs and patients were responsible for at least one onward transmission (i.e. secondary infection) in 49.8% and 50.9% of posterior trees (p = 0.09). However, the Kolmogorov-Smirnov test shows that the cumulative distribution function of secondary infections of HCWs is greater than that of patients (p < 2e-16).

##### Sensitivity analysis

The sensitivity analyses using different values of outlier threshold show different number of imported cases:

- threshold 2 and 3:	4 imported cases
- threshold 4 and 5:	1 imported case

Apart from the number of imported cases, the overall structure of the ancestry reconstruction did not change in those sensitivity analyses (data not shown).

### **eReferences**

1.	Martischang R, Iten A, Arm I, Abbas M, Meyer B, Yerly S, et al. Severe acute respiratory coronavirus virus 2 (SARS-CoV-2) seroconversion and occupational exposure of employees at a Swiss university hospital: A large longitudinal cohort study. Infect Control Hosp Epidemiol. 2021:1-8.

2.	Corman VM, Landt O, Kaiser M, Molenkamp R, Meijer A, Chu DK, et al. Detection of 2019 novel coronavirus (2019-nCoV) by real-time RT-PCR. Euro Surveill. 2020;25(3).

3.	Abbas M, Nunes TR, Cori A, Cordey S, Laubscher F, Baggio S, et al. Explosive nosocomial outbreak of SARS-CoV-2 in a rehabilitation clinic: the limits of genomics for outbreak reconstruction. J Hosp Infect. 2021;Article in press.

4.	Tom J. Petty SC, Ismael Padioleau, Mylène Docquier, Lara Turin, Olivier Preynat-Seauve, Evgeny M. Zdobnov, Laurent Kaiser. Comprehensive Human Virus Screening Using High-Throughput Sequencing with a User-Friendly Representation of Bioinformatics Analysis: a Pilot Study. J Clin Microbiol. 2014;52(9):3351-61.

5.	Fernandes JF, Laubscher F, Held J, Eckerle I, Docquier M, Grobusch MP, et al. Unbiased metagenomic next-generation sequencing of blood from hospitalized febrile children in Gabon. Emerg Microbes Infect. 2020;9(1):1242-4.

6.	Gleizes A, Laubscher F, Guex N, Iseli C, Junier T, Cordey S, et al. Virosaurus A Reference to Explore and Capture Virus Genetic Diversity. Viruses. 2020;12(11).

7.	Kumar S, Stecher G, Li M, Knyaz C, Tamura K. MEGA X: Molecular Evolutionary Genetics Analysis across Computing Platforms. Mol Biol Evol. 2018;35(6):1547-9.

8.	Tamura K. Estimation of the number of nucleotide substitutions when there are strong transition-transversion and G+C-content biase. Molecular Biology and Evolution, Volume 9, Issue 4, Jul 1992, Pages 678–687.

9.	Zou G. A Modified Poisson Regression Approach to Prospective Studies with Binary Data. Am J Epidemiol. 2004;159(7):702–6.

10.	Cori A, Ferguson NM, Fraser C, Cauchemez S. A new framework and software to estimate time-varying reproduction numbers during epidemics. Am J Epidemiol. 2013;178(9):1505-12.

11.	Jombart T, Cori A, Didelot X, Cauchemez S, Fraser C, Ferguson N. Bayesian reconstruction of disease outbreaks by combining epidemiologic and genomic data. PLoS Comput Biol. 2014;10(1):e1003457.

12.	Campbell F, Didelot X, Fitzjohn R, Ferguson N, Cori A, Jombart T. Outbreaker2: a modular platform for outbreak reconstruction. BMC Bioinformatics. 2018;19(Suppl 11).

13.	Sheikh Taslim Ali LW, Eric H. Y. Lau, Xiao-Ke Xu, Zhanwei Du, Ye Wu, Gabriel M. Leung, Benjamin J. Cowling. Serial interval of SARS-CoV-2 was shortened over time by nonpharmaceutical interventions. Science. 2020;369(6507):1106-9.

14.	Bi Q, Wu Y, Mei S, Ye C, Zou X, Zhang Z, et al. Epidemiology and transmission of COVID-19 in 391 cases and 1286 of their close contacts in Shenzhen, China: a retrospective cohort study. The Lancet Infectious Diseases. 2020;20(8):911-9.

15.	Andrew Gelman DBR. Inference from Iterative Simulation Using Multiple Sequences. Statistical Science. 1992;7(4):457-72.

### **eTable 1**. Number of the 285 LTCF employees with ≥ 1 SARS-CoV-2 RT-PCR between March 1^st^ 2020 and June 30, 2020, n (%)

|  | All employees with RT-PCR, n = 106 | Employees with ≥ 1 positive RT-PCR n=36 | Employees with ≥ 1 negative RT-PCR n=70 |
| --- | --- | --- | --- |
| Profession (HCW^1^) | 101 (95) | 35 (97) | 66 (94) |
| Gender (F) | 36 (34) | 26 (72) | 10 (14) |
| Age, median (IQR) | 43 (32 – 52) | 43 (33 – 50) | 43 (32 – 52) |
| Employees included in seroprevalence analysis | 41 (39) | 13 (36) | 28 (40) |
| - Seropositive - Seronegative | 15 (37) 26 (63) | 13 (100)  0 (0) | 2 (7)  26 (93) |
| ^1^ Healthcare workers | | | |

**eFigure 1**. Venn diagram of the number of LTCF employees among all LTCF employees (n=285) who had a SARS-CoV-2 RT-PCR and participated to the seroprevalence survey between March 1st and June 30 2020.

The blue circle represents the number of employees participating to the seroprevalence survey (n=176). The yellow circle represent the number of LTCF employees who underwent SARS-CoV-2 RT-PCR testing (n=106) analyzed in the Laboratory of Virology of Geneva University Hospitals. Forty-one employees participating to the seroprevalence survey had a SARS-CoV-2 PCR testing.

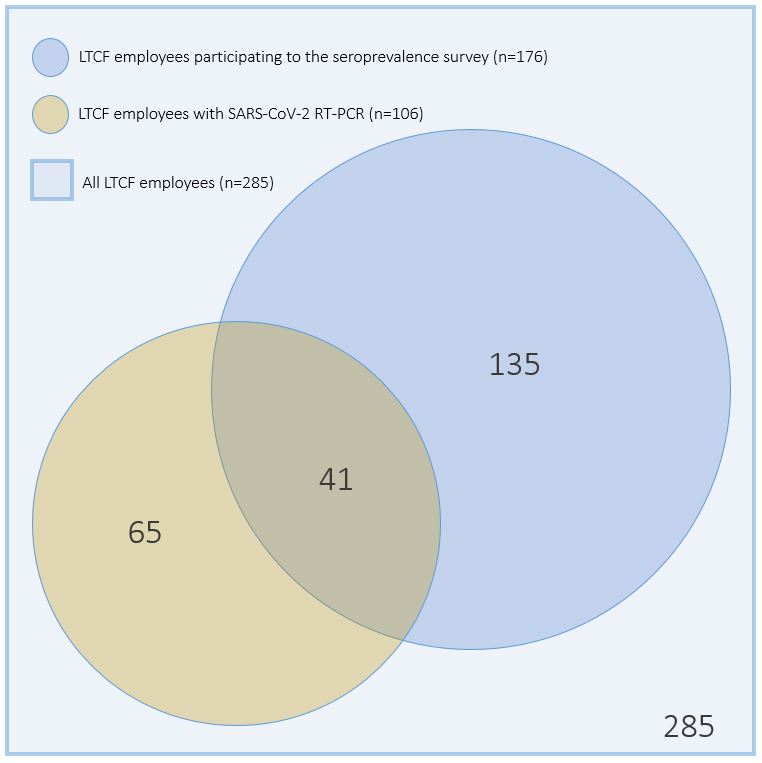

**eFigure 2**. Number of all SARS-CoV-2 RT-PCR performed among LTCF employees and analyzed in the HUG Laboratory of Virology, from March 1 to June 30, 2020.

All SARS-CoV-2 RT-PCR negative results and the first positive result per employee in blue (n=123) and only the first positive SARS-CoV-2 RT-PCR result per employee in red (n=36).

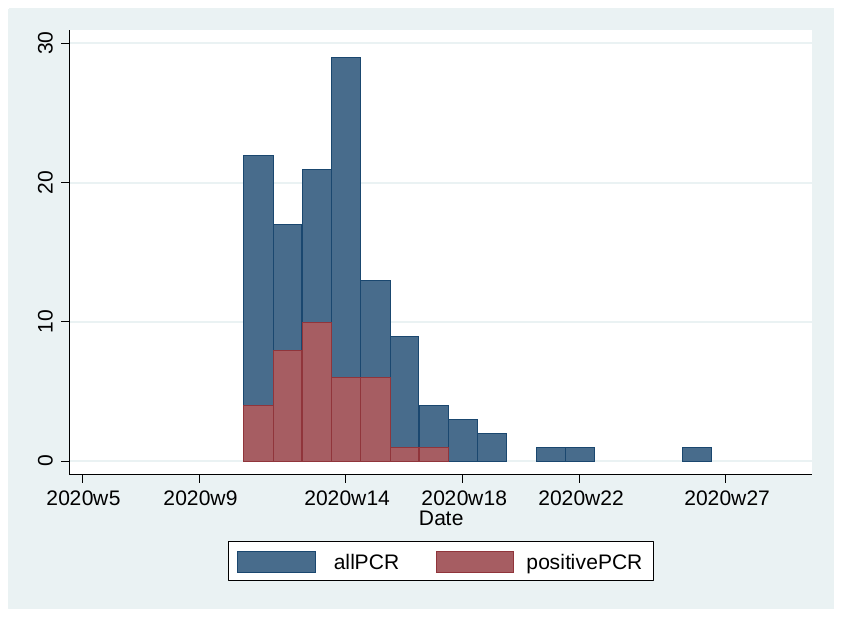

### **eFigure 3**. Flowchart of employees included in the seroprevalence analysis

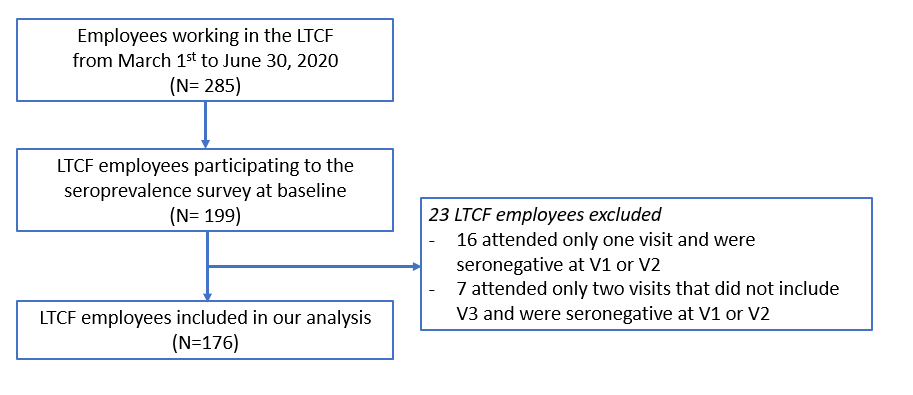

### **eFigure 4**. Ancestry reconstruction.

An ancestry reconstruction summarizing the results of the posterior trees from the outbreaker2 model shows who infected whom with different levels of credibility. All 18 HCWs are represented with the letter “H” and all 10 patients are represented with the letter “C”.

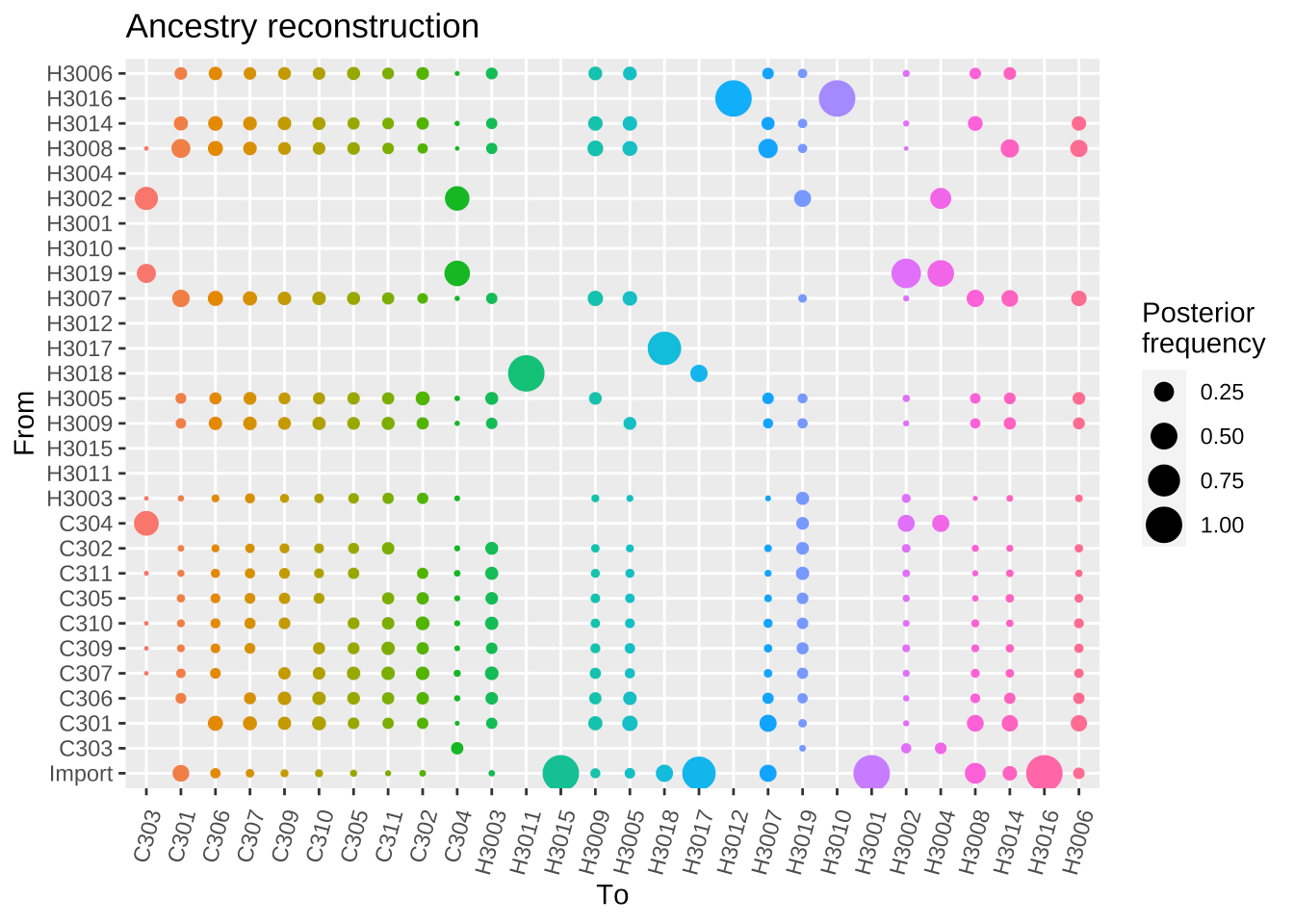

### **eFigure 5**. Maximum posterior transmission tree (displaying only pairs of transmission with a posterior support of > 10%).

HCWs and patients are represented by stethoscope and bed, respectively. Male are in green and female in purple. Each transmission event is represented by an arrow whose color intensity represents the probability of the transmission event, ranging from < 25% (light grey) to 100% (black). Since only the pairs with high posterior support are shown in this figure, it is hard to tell from this which are the imported cases.

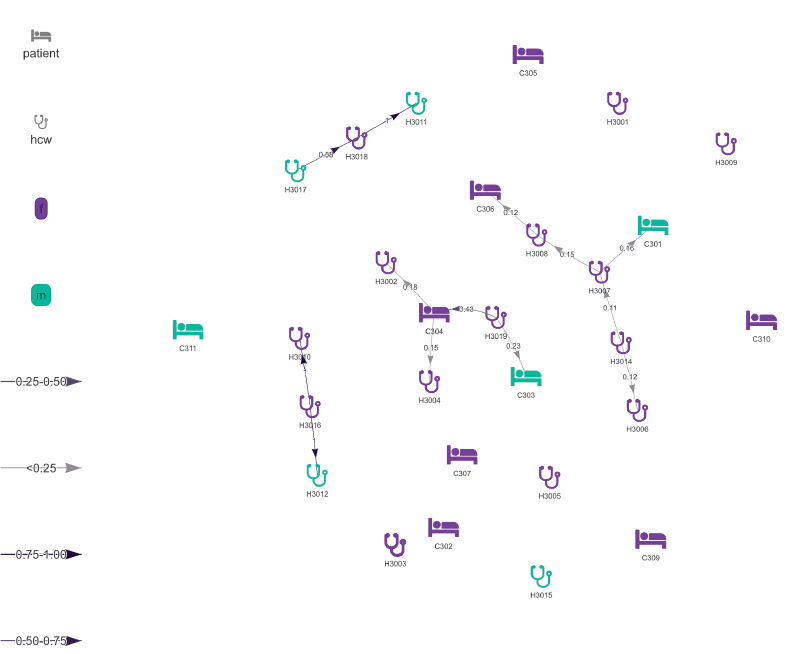

### **eFigure 6**. Number of secondary infections per type of case.

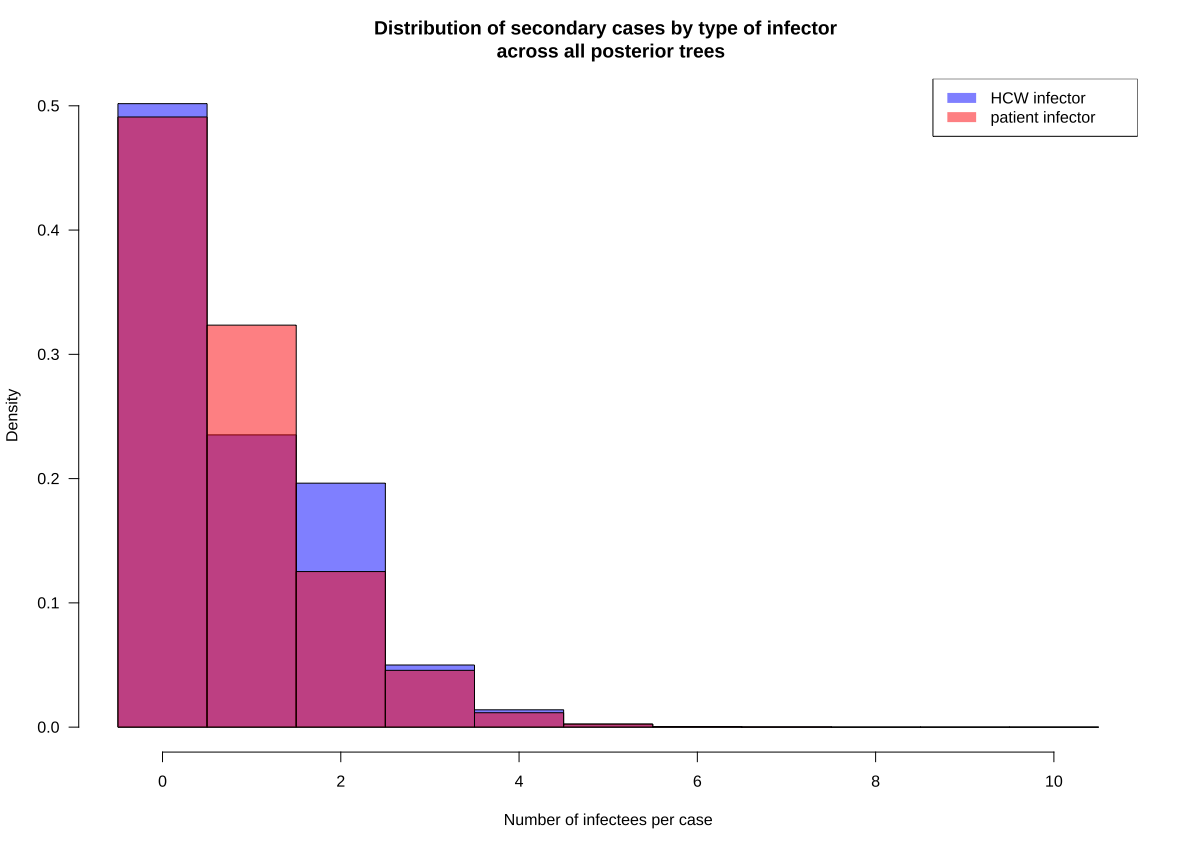
